# Prevalence of SARS-CoV-2 in household members and other close contacts of COVID-19 cases: a serologic study in canton of Vaud, Switzerland

**DOI:** 10.1101/2020.11.27.20239244

**Authors:** Julien Dupraz, Audrey Butty, Olivier Duperrex, Sandrine Estoppey, Vincent Faivre, Julien Thabard, Claire Zuppinger, Gilbert Greub, Giuseppe Pantaleo, Jérôme Pasquier, Valentin Rousson, Malik Egger, Amélie Steiner-Dubuis, Sophie Vassaux, Eric Masserey, Murielle Bochud, Semira Gonseth Nusslé, Valérie D’Acremont

## Abstract

**Background:** Understanding community-based SARS-CoV-2 transmission is crucial to inform public health decisions. Research on SARS-CoV-2 transmission within households and other close settings using serological testing is scarce.

**Methods:** We invited COVID-19 cases diagnosed between February 27 and April 1, 2020 in canton of Vaud, Switzerland, to participate, along with household members and other close contacts. Anti-SARS-CoV-2 IgG antibodies were measured using a Luminex immunoassay. We estimated factors associated with serological status using generalized estimating equations.

**Findings:** Overall, 219 COVID-19 index cases, 302 household members, and 69 other close contacts participated between May 4 and June 27, 2020. More than half of household members (57·2%, 95%CI 49·7-64·3) had developed a serologic response to SARS-CoV-2, while 19·0% (95%CI 10·0-33·2) of other close contacts were seropositive. After adjusting for individual and household characteristics, infection risk was higher in household members aged 65 or more than in younger adults (aOR 3·63, 95%CI 1·05-12·60), and in those not strictly adhering to simple hygiene rules like hand washing (aOR 1·80, 95%CI 1·02-3·17). The risk was lower when more than 5 people outside home were met during the semi-confinement, compared to none (aOR 0·35, 95%CI 0·16-0·74). The individual risk of household members to be seropositive was lower in large households (22% less per each additional person).

**Interpretation:** We find that, during semi-confinement, household members of a COVID-19 case were at very high risk of getting infected, 3 times more than close contacts outside home. This highlights the need to provide clear messages on specific protective measures applicable at home. For elderly couples, who were especially at risk, providing them external support for daily basic activities is essential.

**Funding:** Center for Primary Care and Public Health (Unisanté), Canton of Vaud, Leenaards Foundation, Fondation pour l’Université de Lausanne. SerocoViD is part of Corona Immunitas coordinated by SSPH+.

## Introduction

Tackling a pandemic due to an emerging pathogen, such as the severe acute respiratory syndrome coronavirus 2 (SARS-CoV-2) causing coronavirus disease 2019 (COVID-19), requires early epidemiologic studies to inform public health decisions. The understanding of transmission patterns is especially critical to guide interventions aiming at limiting the occurrence of new cases. In this respect, transmission in promiscuous settings such as households is of particular interest and is at the core of the early investigation protocols provided by the World Health Organisation (WHO Unity Studies) to address the many unknowns related to the COVID-19 pandemic (1,2).

Studies dealing with the transmission of SARS-CoV-2 within households have found secondary attack rates (SAR) ranging from 3·9 to 44·6%, reflecting heterogeneous settings and study designs (3). The evidence regarding transmission to close contacts outside the household tends to show lower SAR (from 0·7 to 5·1%), but attack rates above 50% have been reported in certain circumstances (4–8). Most studies conducted so far are based on the identification of active disease through nucleic acid amplification tests (NAAT), whose sensitivity can be hampered by various factors, such as bad sampling technique, limited sensitivity and sub-optimal timing of specimen collection (9).

The availability of serological assays allows the identification of past infection and thus provides key input into our understanding of the epidemiology of SARS-CoV-2 and its transmission patterns. Nevertheless, studies on SARS-CoV-2 transmission in close settings using serological testing remain scarce. So far, most of them found SAR close to 35% within households (10–13). However, with the exception of a nationwide Spanish study, their estimations are based on less than 80 household members, and none of them includes a thorough investigation of factors associated with seropositivity. Regarding close contacts outside the household, research shows SAR ranging from 0 to 13·7%, but study designs and settings are disparate (12,14–16). Furthermore, the amount of available serological assays is quickly growing, often with limited external validation of their accuracy, and concerns are emerging regarding their accuracy in the setting of seroepidemiological studies because of the lower median level of antibodies in participants compared to clinical studies (17).

This work was part of *SerocoViD*, a community-based seroepidemiological study of SARS-CoV-2 infection conducted in canton of Vaud, Switzerland, embedded within a nationwide program, Corona Immunitas (18). Taking advantage of prior development and validation of a highly sensitive serological assay carried out locally (19), the objective was to determine the prevalence of anti-SARS-CoV-2 IgG antibodies among household members and other close contacts of index COVID-19 cases, and to identify factors associated with seropositivity in these highly exposed people.

## Methods

### Study design and participants

*SerocoViD* is a cross-sectional community-based seroepidemiological study of SARS-CoV-2 infection conducted in canton of Vaud (French-speaking region of Switzerland, 806’088 inhabitants on December 31, 2019). The study was launched at the end of April 2020, coinciding with the easing of semi-confinement measures taken in Switzerland in mid-March. The Cantonal Ethics Committee of Vaud, Switzerland (ID 2020-00887) approved the protocol. We used reporting guidelines for observational studies developed by the STROBE initiative. <u>The full protocol is available online.</u>

From February 27 (first confirmed case in canton of Vaud) to March 4, 2020, all COVID-19 cases underwent contact tracing by local authorities. At that time, a close contact was any individual who had been within two metres of an infected person for at least 15 minutes, starting 24 hours before illness onset. Given the exponential growth of the number of cases, contact tracing was stopped from week 2 of the epidemic. For the same reason, from March 9, 2020, diagnostic testing was limited to healthcare personal, hospitalised people and individuals at increased risk for severe illness because of their age or comorbidities in the entire country.

We sampled confirmed COVID-19 cases from the cantonal registry (total n≈3’700). With the exception of three people (one deceased, two who returned home abroad), all confirmed cases from week 1 were invited to participate in the study (n=13), along with their close contacts identified by contact tracing (n=117). Additionally, all cases aged between 6 months and 19 years (n=66) and a random sample of non-institutionalised cases aged 20 and more (n=368) who were tested positive during weeks 2 to 5 (from March 5 to April 1, 2020) were invited to take part in the study. In order to extend the age range of confirmed cases for whom a contact tracing procedure had been performed, the study team conducted complementary tracing procedures for three adolescent cases, thus identifying 20 additional close contacts outside the household.

Overall, this resulted in the solicitation of 447 confirmed cases (called thereafter index cases) and 137 close contacts not belonging to the household of index cases. Moreover, index case participants were asked to invite all their household members aged 6 months or more to take part in the study.

### Procedures

Index cases and their close contacts identified by contact tracing were invited by letters. Participants registered into the study and answered the study questionnaire (available in French and English) via an online platform. Those having technical or understanding difficulties could call a dedicated hotline to request assistance or get help from the study team during the study visit. The questionnaire covered the following topics: socio-demographic information, medical history, history of symptoms compatible with COVID-19 and use of health services, living conditions and household characteristics, contacts with other people in private and professional settings, and compliance with measures aimed at controlling the epidemic. The full questionnaire is available as supplementary material.

Study visits took place in four centres distributed over the cantonal territory between May 4 and June 27, 2020. A venous blood sample was collected to proceed with serological testing. We offered a home visit by a mobile study team to people at increased risk for severe illness from COVID-19 due to age or comorbidities. All participants (or their legal representative) provided written informed consent.

### Detection of anti-SARS-CoV-2 antibodies

We measured anti-SARS-CoV-2 IgG antibodies targeting the spike (S) protein in its native trimeric form using a Luminex immunoassay. This test was developed by the Lausanne University Hospital, Switzerland, in collaboration with the École Polytechnique Fédérale de Lausanne (EPFL), and compared with five commercially available immunoassays detecting IgG against the N protein and the monomeric moieties of the S1 protein (19). The in-house Luminex S protein trimer IgG assay was 99·2% specific in sera from people infected with pre-pandemic coronaviruses or from patients with autoimmune diseases, and proved to be more sensitive (96·7%) than commercial tests in hospitalized patients with moderate to severe disease 16 to 33 days post-symptoms. The threshold for a positive result was defined at an antibody Multiplex Fluorescent Immunoassay (MFI) ratio of ≥6, and a negative result for ratios <6.

### Statistical analysis

We calculated the proportion of index cases with a positive serology test result and computed a 95% confidence interval using the Clopper-Pearson method. Significant clustering of infections within households has been reported in previous research (20). In order to account for correlation between close contacts of a same index case, we used generalised estimating equations (GEE) with an exchangeable correlation structure to estimate the seroprevalence and corresponding 95% confidence interval among contacts. Odds ratios (OR) were computed to measure the strength of the association between each independent variable and serology test result. We used GEE to account for correlation between contacts of a same index case and calculated OR with their 95% confidence interval and p-value using a logit link function. Finally, a multivariable regression model using GEE was fitted to measure the adjusted association of individual and household characteristics with serology test result among household members. The variable “number of chronic medical conditions”, that was not available for children and teens, was not included in the model. Considering the potential influence of past diagnostic testing for SARS-CoV-2 on the reporting of symptoms, we proceeded to a sensitivity analysis among contacts not reporting previous nasal or throat swabbing. We performed statistical analysis using Stata/IC version 16.1. There was no imputation of missing values.

## Results

Two-hundred and nineteen index cases (49·0%), aged 2 to 90 years (mean 48·7, SD 19·3), participated in the study, of which 55·7% considered themselves as women. They reported 421 household members, of which 302 (71·7%), aged 1 to 87 years (mean 37·0, SD 21·3), took part in the study. Sixty-nine (50·4%) close contacts outside the household, aged 9 to 85 years (mean 47·8, SD 17·0) participated.

### Prevalence of seropositivity in the different groups

Most index cases (215/219, 98·2%) had a positive serological test result (95%CI 95·4-99·5; **figure 1**). The crude proportion of positives was 53·0% in household members (160/302) and 17·4% among close contacts outside the household (12/69). When taking into account correlation, the seroprevalence was 57·2% in household members (95%CI 49·7-64·3) and 19·0% in close contacts outside the household (95%CI 10·0-33·2).

**Figure 1:**
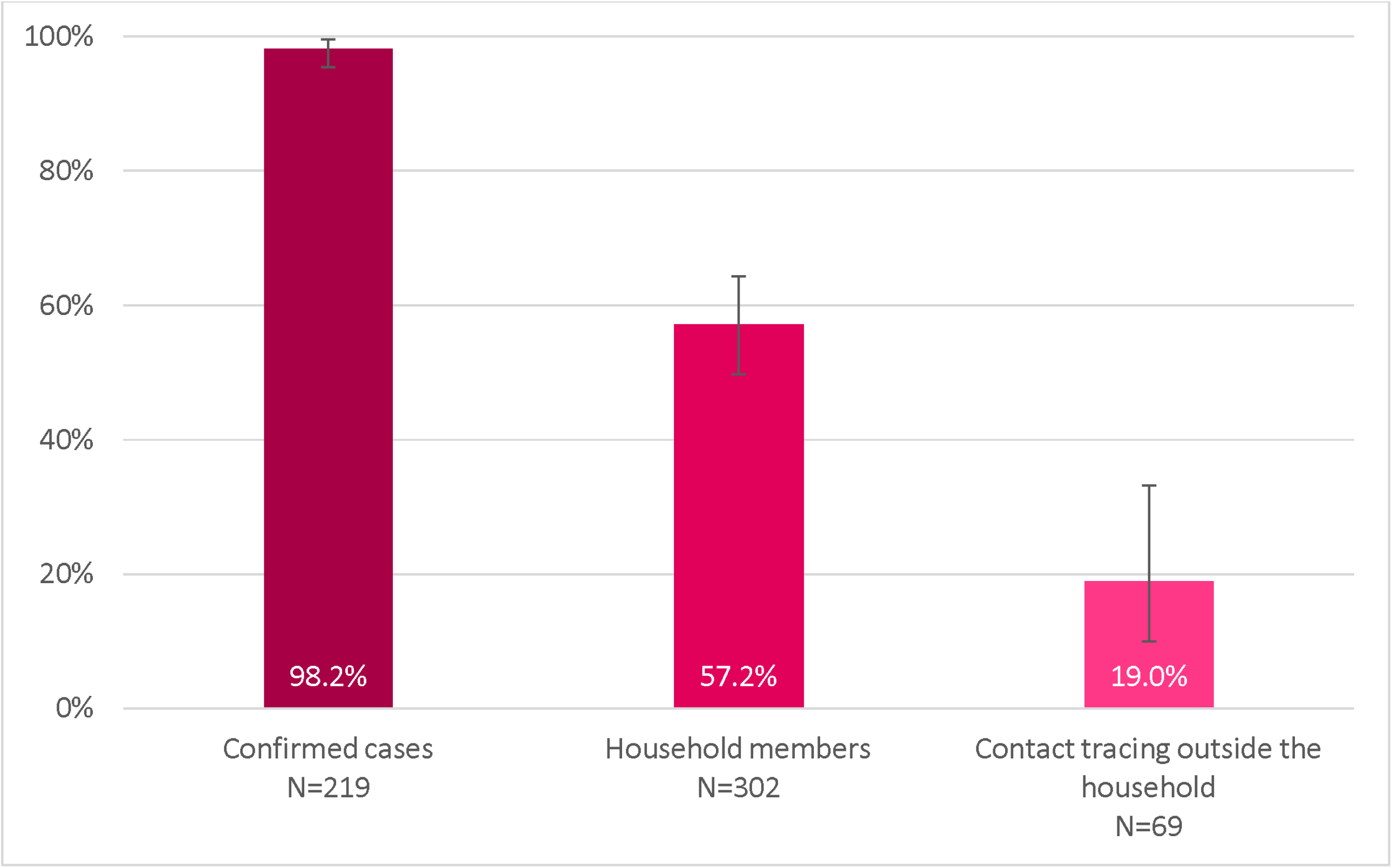
Percentage of participants with a positive serology test result, by type of participant. Index cases: crude proportion, calculation of 95% confidence interval using the Clopper-Pearson method. Household members and close contacts outside the household: proportion and corresponding 95% confidence interval estimated using GEE (exchangeable correlation structure).

### Association of general characteristics and medical history of contacts with seropositivity

A higher proportion of household members aged 65 to 75 (85·7%) and 75 or more (83·3%) were seropositive (**table 1**). Among close contacts identified by contact tracing, no participant aged less than 20 was seropositive. No association between serological test result and gender or level of education was found (**figure 2**). Household members currently smoking had lower odds of infection than non-smokers (OR 0·56, 95%CI 0·32-0·96). In close contacts outside the household, seroprevalence was 30·3% and 5·7% in overweight/obese and normal/underweight participants respectively. The association between weight and serological status was weaker among household members, but in this group, adult participants suffering from one or more chronic conditions (outside overweight/obesity) had higher odds of infection (OR 1·94, 95%CI 1·04-3·62).

**Table 1:** Serology test result according to general characteristics and medical history, stratified by type of participant.

|  | Index cases (N=219) |  | Household members (N=302) |  |  | Close contacts outside the household (N=69) |  |  |
| --- | --- | --- | --- | --- | --- | --- | --- | --- |
|  | n (%) seropositive |  | n (%) seropositive | Odds ratio [95% CI] | p-value | n (%) seropositive | Odds ratio [95% CI] | p-value |
| **All participants** | 215/219 (98.2) |  | 160/302 (53.0) |  |  | 12/69 (17.4) |  |  |
| Age |  |  |  |  | 0.119 |  |  | 0.928 |
| 6mo-<5y | 1/2 (50.0) |  | 5/11 (45.5) | 0.92 [0.37-2.29] |  | no participant | .. |  |
| 5y-<10y | 1/1 (100.0) |  | 12/22 (54.6) | 1.16 [0.55-2.44] |  | 0/1 (0.0) | .. |  |
| 10y-<15y | 2/2 (100.0) |  | 15/32 (46.9) | 0.93 [0.49-1.75] |  | 0/3 (0.0) | .. |  |
| 15y-<20y | 20/21 (95.2) |  | 9/19 (47.4) | 1.17 [0.56-2.45] |  | 0/2 (0.0) | .. |  |
| 20y-<40y | 42/43 (97.7) |  | 37/76 (48.7) | reference |  | 3/15 (20.0) | reference |  |
| 40y-<65y | 103/104 (99.0) |  | 60/116 (51.7) | 0.83 [0.52-1.30] |  | 7/38 (18.4) | 0.92 [0.22-3.87] |  |
| 65y-<75y | 31/31 (100.0) |  | 12/14 (85.7) | 3.98 [1.03-15.44] |  | 2/9 (22.2) | 1.32 [0.16-11.00] |  |
| 75y or more | 15/15 (100.0) |  | 10/12 (83.3) | 5.25 [1.16-23.72] |  | 0/1 (0.0) | .. |  |
| Gender |  |  |  |  | 0.164 |  |  | 0.124 |
| male | 94/96 (97.9) |  | 74/146 (50.7) | reference |  | 8/30 (26.7) | reference |  |
| female | 120/122 (98.4) |  | 86/156 (55.1) | 1.27 [0.91-1.76] |  | 4/39 (10.3) | 0.36 [0.10-1.32] |  |
| other | 1/1 (100.0) |  | no participant | .. |  | no participant | .. |  |
| Current smoker * |  |  |  |  | 0.034 |  |  | 0.825 |
| no | 191/194 (98.5) |  | 142/256 (55.5) | reference |  | 10/59 (17.0) | reference |  |
| yes | 23/24 (95.8) |  | 18/46 (39.1) | 0.56 [0.32-0.96] |  | 2/9 (22.2) | 1.21 [0.22-6.53] |  |
| Weight status |  |  |  |  | 0.146 |  |  | 0.011 |
| normal or underweight | 104/106 (98.1) |  | 92/184 (50.0) | reference |  | 2/35 (5.7) | reference |  |
| overweight or obese | 110/112 (98.2) |  | 66/111 (59.5) | 1.37 [0.90-2.09] |  | 10/33 (30.3) | 6.74 [1.54-29.50] |  |
| **Adult participants only** |  |  |  |  |  |  |  |  |
| Education |  |  |  |  | 0.196 |  |  | 0.423 |
| lower secondary or less | 21/21 (100.0) |  | 21/33 (63.6) | reference |  | 3/10 (30.0) | reference |  |
| upper secondary | 58/59 (98.3) |  | 46/82 (56.1) | 0.93 [0.46-1.90] |  | 5/23 (21.7) | 0.64 [0.13-3.21] |  |
| tertiary | 108/109 (99.1) |  | 51/100 (51.0) | 0.61 [0.30-1.24] |  | 4/30 (13.3) | 0.33 [0.06-1.83] |  |
| Chronic medical conditions † |  |  |  |  | 0.037 |  |  | 0.148 |
| none | 131/133 (98.5) |  | 86/169 (50.9) | reference |  | 6/44 (13.6) | reference |  |
| one or more | 58/58 (100.0) |  | 32/47 (68.1) | 1.94 [1.04-3.62] |  | 6/19 (31.6) | 2.48 [0.73-8.48] |  |
| Hypertension |  |  |  |  | 0.110 |  |  | 0.034 |
| no | 152/154 (98.7) |  | 95/182 (52.2) | reference |  | 7/50 (14.0) | reference |  |
| yes | 36/36 (100.0) |  | 21/31 (67.7) | 1.81 [0.87-3.74] |  | 5/11 (45.5) | 4.48 [1.12-18.01] |  |
| Diabetes |  |  |  |  | 0.489 |  |  | 0.082 |
| no | 169/171 (98.8) |  | 111/206 (53.9) | reference |  | 10/57 (17.5) | reference |  |
| yes | 15/15 (100.0) |  | 4/6 (66.7) | 1.75 [0.36-8.48] |  | 2/3 (66.7) | 8.59 [0.76-96.92] |  |
| Cardiovascular disease |  |  |  |  | 0.239 |  |  | .. |
| no | 170/172 (98.8) |  | 103/196 (52.6) | reference |  | 12/58 (20.7) | .. |  |
| yes | 12/12 (100.0) |  | 9/12 (75.0) | 2.03 [0.62-6.62] |  | 0/2 (0.0) | .. |  |
| Kidney disease |  |  |  |  | .. |  |  | .. |
| no | 181/183 (98.9) |  | 115/212 (54.3) | .. |  | 12/60 (20.0) | .. |  |
| yes | 3/3 (100.0) |  | no participant | .. |  | 0/1 (0.0) | .. |  |
| Chronic respiratory disease |  |  |  |  | 0.196 |  |  | .. |
| no | 177/179 (98.9) |  | 106/202 (52.5) | reference |  | 12/59 (20.3) | .. |  |
| yes | 7/7 (100.0) |  | 4/5 (80.0) | 3.79 [0.50-28.52] |  | 0/2 (0.0) | .. |  |
| Immunodeficiency |  |  |  |  | 0.596 |  |  | .. |
| no | 174/176 (98.9) |  | 110/202 (54.5) | reference |  | 12/58 (20.7) | .. |  |
| yes | 12/12 (100.0) |  | 4/9 (44.4) | 0.74 [0.25-2.23] |  | 0/3 (0.0) | .. |  |
| Cancer |  |  |  |  | .. |  |  | .. |
| no | 177/179 (98.9) |  | 113/209 (54.1) | .. |  | 12/58 (20.7) | .. |  |
| yes | 4/4 (100.0) |  | no participant | .. |  | 0/2 (0.0) | .. |  |
| Other chronic condition |  |  |  |  | 0.325 |  |  | .. |
| no | 159/160 (99.4) |  | 97/184 (52.7) | reference |  | 12/55 (21.8) | .. |  |
| yes | 24/25 (96.0) |  | 18/28 (64.3) | 1.49 [0.68-3.27] |  | 0/7 (0.0) | .. |  |

Calculation of odds ratio and p-value: correlation between close contacts of a same index case taken into account using GEE (exchangeable correlation structure, logit link function). \* Children aged less than 12 considered non-smokers. † Among all following conditions, except "other chronic condition".

**Figure 2:**
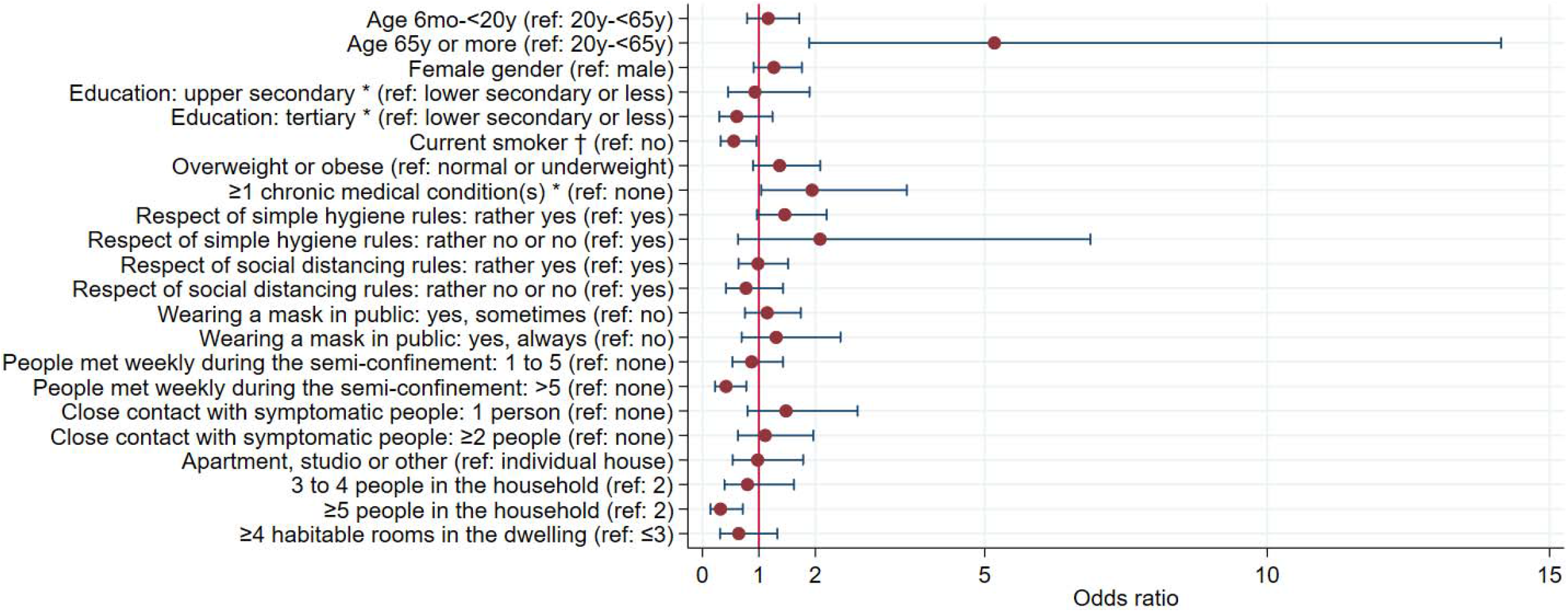
Association between characteristics of household members of index cases and seropositivity (bivariate analysis) Calculation of odds ratio: correlation between household members of a same index case taken into account using GEE (exchangeable correlation structure, logit link function). Error bars represent the limits of the 95% confidence interval for the odds ratio. * Adult participants only. † Children aged less than 12 considered non-smokers.

### Association of adherence to protective measures, contacts with other people, and living conditions with seropositivity

Close contacts not strictly adhering to simple hygiene rules tended to have higher odds of infection (**table 2, figure 2**). We found no association between serology and the compliance with social distancing rules. Interestingly, close contacts outside the household who reported always wearing a mask in public tended to be more frequently seropositive (OR 4·36, 95%CI 1·06-17·83). Positive test results were less frequent in household members who had met more than five people per week during the semi-confinement compared to none (OR 0·42, 95%CI 0·22-0·78), but there was no association with the number of close encounters with symptomatic individuals. Seroprevalence significantly decreased with increasing household size. We found that 66·1% of participants living with one other person only (the index COVID-19 case) had a positive test result, contrasting with participants living with five people or more, who showed a 26·0% risk to be seropositive (OR 0·19, 95%CI 0·06-0·62). There was an inverse relationship between household size and mean age of participants in the household (**supplementary table 1**).

**Table 2:** Serology test result according to adherence to measures aimed at decreasing transmission, contacts with other people, and living conditions, stratified by type of participant.

|  | Household members (N=302) |  |  | Close contacts outside the household (N=69) |  |  |
| --- | --- | --- | --- | --- | --- | --- |
|  | n (%) seropositive | Odds ratio [95% CI] | p-value | n (%) seropositive | Odds ratio [95% CI] | p-value |
| **<u>Respect of measures and contacts with other people</u>** |  |  |  |  |  |  |
| Respect of simple hygiene rules (washing hands regularly, sneezing into the elbow, etc.) |  |  | 0.123 |  |  | 0.272 |
| yes | 114/215 (53.0) | reference |  | 8/56 (14.3) | reference |  |
| rather yes | 40/74 (54.1) | 1.46 [0.97-2.20] |  | 4/13 (30.8) | 2.22 [0.54-9.21] |  |
| rather no or no | 4/9 (44.4) | 2.08 [0.63-6.87] |  | no participant | .. |  |
| Respect of social distancing rules (physical distancing, avoid shaking hands or kissing, etc.) |  |  | 0.868 |  |  | 0.114 |
| yes | 87/157 (55.4) | reference |  | 7/53 (13.2) | reference |  |
| rather yes | 56/105 (53.3) | 0.99 [0.64-1.52] |  | 5/15 (33.3) | 2.94 [0.77-11.23] |  |
| rather no or no | 15/36 (41.7) | 0.77 [0.42-1.43] |  | 0/1 (0.0) | .. |  |
| Wearing a mask in public |  |  | 0.657 |  |  | 0.093 |
| no | 79/163 (48.5) | reference |  | 5/38 (13.2) | reference |  |
| yes, sometimes | 54/98 (55.1) | 1.15 [0.75-1.74] |  | 2/17 (11.8) | 1.06 [0.21-5.35] |  |
| yes, always | 27/40 (67.5) | 1.31 [0.70-2.45] |  | 5/13 (38.5) | 4.36 [1.06-17.83] |  |
| Weekly number of people met outside home during the semi-confinement |  |  | 0.009 |  |  | 0.761 |
| none | 65/100 (65.0) | reference |  | 2/17 (11.8) | reference |  |
| 1 to 5 | 73/140 (52.1) | 0.87 [0.53-1.43] |  | 6/34 (17.7) | 1.26 [0.25-6.35] |  |
| more than 5 | 21/61 (34.4) | 0.42 [0.22-0.78] |  | 4/17 (23.5) | 1.86 [0.32-10.75] |  |
| Close contact with people outside home having symptoms suggestive of COVID-19 |  |  | 0.456 |  |  | 0.099 |
| none | 127/233 (54.5) | reference |  | 2/14 (14.3) | reference |  |
| 1 person | 17/34 (50.0) | 1.48 [0.80-2.75] |  | 5/42 (11.9) | 0.88 [0.15-5.01] |  |
| 2 people or more | 16/35 (45.7) | 1.11 [0.63-1.96] |  | 5/12 (41.7) | 4.36 [0.68-27.99] |  |
| **<u>Living conditions and household characteristics \*</u>** |  |  |  |  |  |  |
| Housing type † |  |  | 0.946 |  |  |  |
| individual house | 83/158 (52.5) | reference |  |  |  |  |
| apartment, studio or other | 77/144 (53.5) | 0.98 [0.54-1.79] |  |  |  |  |
| Number of people in the household † |  |  | 0.046 |  |  |  |
| 2 | 39/59 (66.1) | reference |  |  |  |  |
| 3 | 31/51 (60.8) | 0.82 [0.35-1.93] |  |  |  |  |
| 4 | 44/74 (59.5) | 0.78 [0.34-1.77] |  |  |  |  |
| 5 | 33/68 (48.5) | 0.43 [0.17-1.06] |  |  |  |  |
| 6 or more | 13/50 (26.0) | 0.19 [0.06-0.62] |  |  |  |  |
| Number of habitable rooms (besides kitchen) in the dwelling ‡ |  |  | 0.441 |  |  |  |
| 3 or less | 35/54 (64.8) | reference |  |  |  |  |
| 4 to 6 | 96/183 (52.5) | 0.67 [0.32-1.40] |  |  |  |  |
| 7 or more | 29/65 (44.6) | 0.55 [0.21-1.47] |  |  |  |  |

All participants, including children and teens. Calculation of odds ratio and p-value: correlation between close contacts of a same index case taken into account using GEE (exchangeable correlation structure, logit link function). \* Not relevant for close contacts outside household. † Answer of the index case taken for all household members. ‡ Answer of the index case taken for all household members, except two households where information from index case was missing (mean of answers reported by other household members taken instead).

We finally estimated the adjusted association of individual and household characteristics with serology test result among household members (**table 3**). Given the association between household size and mean age of participating household members, this variable was included in the model. The odds of infection was almost four times higher in household members aged 65 or more than in the younger age group (adjusted OR 3·63, 95%CI 1·05-12·60). The association of current smoking with negative serology observed in bivariable analysis faded in multivariable model (aOR 0·73, 95%CI 0·38-1·39). Although overweight/obesity tended to be associated with higher odds of infection, this association was not statistically significant at the 0.05 level. In comparison with bivariable analysis, we observed a strengthening of the relation between the absence of strict adherence to simple hygiene rules and positive serology testing (aOR 1·80, 95%CI 1·02-3·17). However, there was no indication of a link with adherence to social distancing rules or mask wearing. The association of a greater number of social contacts during the semi-confinement with lower odds of infection was confirmed in multivariable analysis (aOR 0·35, 95%CI 0·16-0·74). On the other hand, close encounters with symptomatic individuals tended to be associated with positive serology, but this tendency was not statistically significant at the 0.05 level. Household characteristics did not show a significant association with serological test result. Inclusion of the mean age of household members in the model slightly weakened the association of the number of people in the household with the outcome (without mean age variable, aOR 0·76, 95%CI 0·59-0·99, p-value 0·039). Adding characteristics of the index case to the model (age, gender) yielded comparable estimates (results not shown).

**Table 3:** Association of individual and household characteristics with serology test result among household members.

|  | Adjusted OR [95% CI] | p-value |
| --- | --- | --- |
| **<u>Characteristics of household member</u>** |  |  |
| Age (ref: 20y-<65y) |  |  |
| 6mo-<20y | 0.92 [0.54-1.59] | 0.775 |
| 65y or more | 3.63 [1.05-12.60] | 0.042 |
| Gender (ref: male) |  |  |
| female | 1.37 [0.90-2.08] | 0.137 |
| Current smoker (ref: no) |  |  |
| yes | 0.73 [0.38-1.39] | 0.339 |
| Weight status (ref: normal or underweight) |  |  |
| overweight or obese | 1.48 [0.90-2.43] | 0.125 |
| Respect of simple hygiene rules (washing hands regularly, sneezing into the elbow, etc.) (ref: yes) |  |  |
| rather yes, rather no, or no | 1.80 [1.02-3.17] | 0.041 |
| Respect of social distancing rules (physical distancing, avoid shaking hands or kissing, etc.) (ref: yes) |  |  |
| rather yes, rather no, or no | 1.06 [0.62-1.82] | 0.831 |
| Wearing a mask in public (ref: no) |  |  |
| yes, sometimes | 1.02 [0.61-1.72] | 0.926 |
| yes, always | 0.94 [0.43-2.09] | 0.885 |
| Weekly number of people met outside home during the semi-confinement (ref: 0) |  |  |
| 1 to 5 | 0.70 [0.40-1.21] | 0.201 |
| more than 5 | 0.35 [0.16-0.74] | 0.006 |
| Close contact with people outside home having symptoms suggestive of COVID-19 (ref: none) |  |  |
| 1 person | 1.29 [0.62-2.67] | 0.495 |
| 2 people or more | 1.72 [0.86-3.45] | 0.125 |
| **<u>Characteristics of household</u>** |  |  |
| Highest education level among adult household members (ref: lower secondary or less) |  |  |
| upper secondary | 1.08 [0.21-5.54] | 0.922 |
| tertiary | 1.64 [0.34-7.95] | 0.541 |
| Housing type † (ref: individual house) |  |  |
| apartment, studio or other | 0.87 [0.39-1.95] | 0.738 |
| Number of people in the household † |  |  |
| one-person increase | 0.78 [0.56-1.08] | 0.135 |
| Number of habitable rooms (besides kitchen) in the dwelling ‡ |  |  |
| one-room increase | 0.98 [0.76-1.25] | 0.843 |
| Mean age of participating household members |  |  |
| one-year increase | 1.00 [0.97-1.04] | 0.793 |

Multivariable regression model; 291/302 household members included in model. Within-household correlation taken into account using GEE (exchangeable correlation structure, logit link function). \* Children aged less than 12 considered non-smokers. † Answer of the index case taken for all household members. ‡ Answer of the index case taken for all household members, except two households where information from index case was missing (mean of answers reported by other household members taken instead).

### Prevalence and clinical presentation of flu-like episodes, and use of health services

The occurrence of one flu-like episode or more since the end of February 2020 was strongly associated with positive serological testing, both in household members (OR 3·55, 95%CI 2·37-5·32, **table 4**) and close contacts outside the household (OR 8·64, 95%CI 1·77-42·12). The proportion of asymptomatic seropositive individuals (i.e. not reporting any flu-like episode) was 21·4% in household members, and 16·7% in close contacts outside the household. With the exception of chest pain, all reported symptoms were associated with a positive serology. This was particularly evident in household members mentioning new-onset anosmia or ageusia, of which 92·8% were seropositive (OR 6·24, 95%CI 3·46-11·24). When limiting the analysis to participants not reporting previous nasal or throat swabbing, the strength of the association between symptoms and serology generally increased (**supplementary table 2**). Half the seropositive household members not mentioning prior PCR testing reported tiredness (49·6%; **figure 3**), followed by headache (44·1%), cough (37·1%), fever (36·8%), aching muscle or joints (36·6%), and anosmia or ageusia (35·9%). Gastrointestinal symptoms were infrequent. Half of seropositive household members (46·3%) reported contact with a medical provider, and 6·3% were hospitalised. Figures were comparable among seropositive close contacts outside the household (41·7% and 8·3%, respectively). However, hospitalisation rate was higher in index cases (14·7%).

**Table 4:**
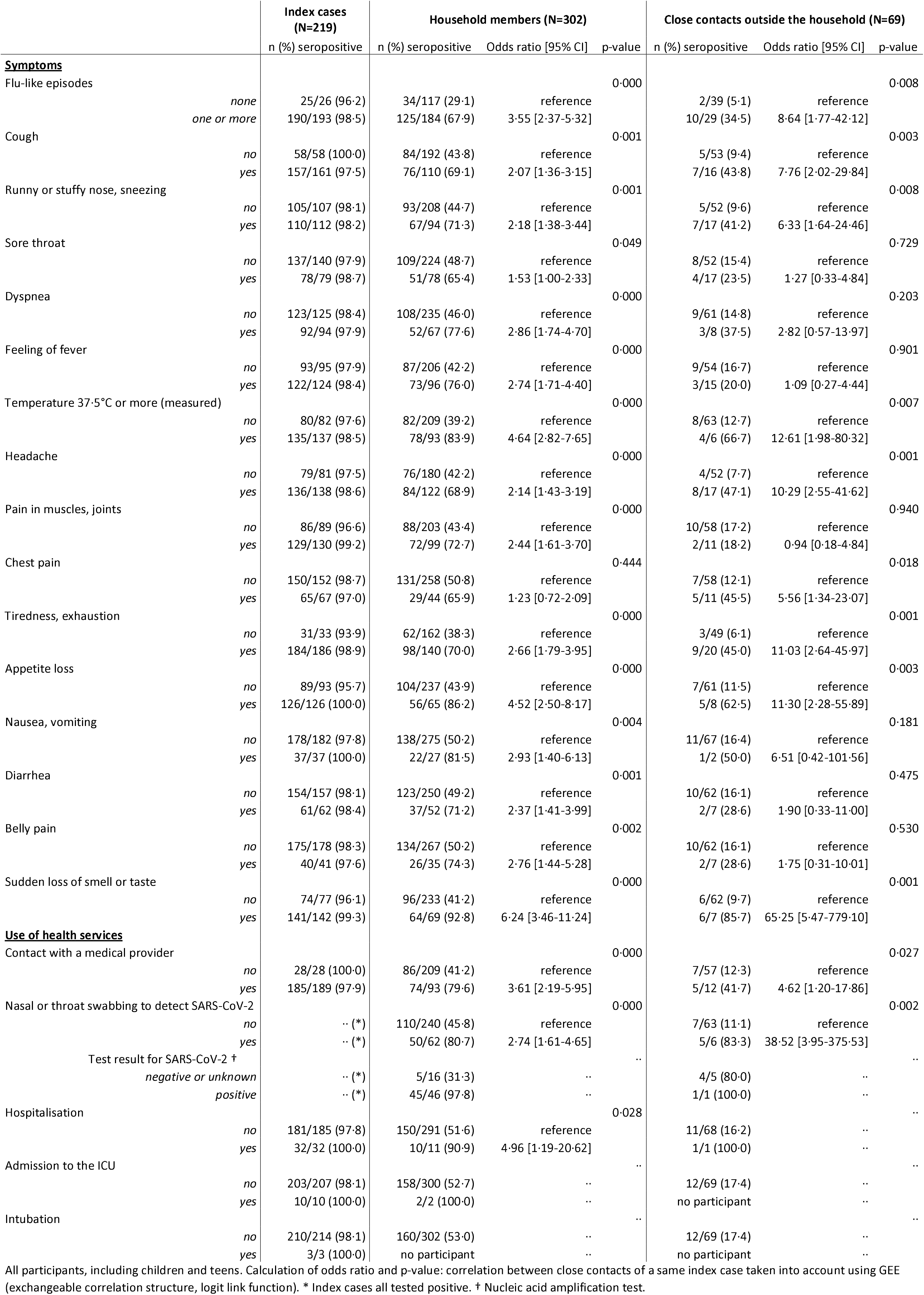
Serology test result according to symptoms and use of health services since the end of February 2020, stratified by type of participant.

|  |  | Index cases<br>(N=219) | Household members (N=302) |  |  | Close contacts outside the household (N=69) |  |  |
| --- | --- | --- | --- | --- | --- | --- | --- | --- |
|  |  | n (%) seropositive | n (%) seropositive | Odds ratio [95% CI] | p-value | n (%) seropositive | Odds ratio [95% CI] | p-value |
| **Symptoms** |  |  |  |  |  |  |  |  |
| Flu-like episodes |  |  |  |  | 0.000 |  |  | 0.008 |
|  | *none* | 25/26 (96.2) | 34/117 (29.1) | reference |  | 2/39 (5.1) | reference |  |
|  | *one or more* | 190/193 (98.5) | 125/184 (67.9) | 3.55 [2.37-5.32] |  | 10/29 (34.5) | 8.64 [1.77-42.12] |  |
| Cough |  |  |  |  | 0.001 |  |  | 0.003 |
|  | *no* | 58/58 (100.0) | 84/192 (43.8) | reference |  | 5/53 (9.4) | reference |  |
|  | *yes* | 157/161 (97.5) | 76/110 (69.1) | 2.07 [1.36-3.15] |  | 7/16 (43.8) | 7.76 [2.02-29.84] |  |
| Runny or stuffy nose, sneezing |  |  |  |  | 0.001 |  |  | 0.008 |
|  | *no* | 105/107 (98.1) | 93/208 (44.7) | reference |  | 5/52 (9.6) | reference |  |
|  | *yes* | 110/112 (98.2) | 67/94 (71.3) | 2.18 [1.38-3.44] |  | 7/17 (41.2) | 6.33 [1.64-24.46] |  |
| Sore throat |  |  |  |  | 0.049 |  |  | 0.729 |
|  | *no* | 137/140 (97.9) | 109/224 (48.7) | reference |  | 8/52 (15.4) | reference |  |
|  | *yes* | 78/79 (98.7) | 51/78 (65.4) | 1.53 [1.00-2.33] |  | 4/17 (23.5) | 1.27 [0.33-4.84] |  |
| Dyspnea |  |  |  |  | 0.000 |  |  | 0.203 |
|  | *no* | 123/125 (98.4) | 108/235 (46.0) | reference |  | 9/61 (14.8) | reference |  |
|  | *yes* | 92/94 (97.9) | 52/67 (77.6) | 2.86 [1.74-4.70] |  | 3/8 (37.5) | 2.82 [0.57-13.97] |  |
| Feeling of fever |  |  |  |  | 0.000 |  |  | 0.901 |
|  | *no* | 93/95 (97.9) | 87/206 (42.2) | reference |  | 9/54 (16.7) | reference |  |
|  | *yes* | 122/124 (98.4) | 73/96 (76.0) | 2.74 [1.71-4.40] |  | 3/15 (20.0) | 1.09 [0.27-4.44] |  |
| Temperature 37.5°C or more (measured) |  |  |  |  | 0.000 |  |  | 0.007 |
|  | *no* | 80/82 (97.6) | 82/209 (39.2) | reference |  | 8/63 (12.7) | reference |  |
|  | *yes* | 135/137 (98.5) | 78/93 (83.9) | 4.64 [2.82-7.65] |  | 4/6 (66.7) | 12.61 [1.98-80.32] |  |
| Headache |  |  |  |  | 0.000 |  |  | 0.001 |
|  | *no* | 79/81 (97.5) | 76/180 (42.2) | reference |  | 4/52 (7.7) | reference |  |
|  | *yes* | 136/138 (98.6) | 84/122 (68.9) | 2.14 [1.43-3.19] |  | 8/17 (47.1) | 10.29 [2.55-41.62] |  |
| Pain in muscles, joints |  |  |  |  | 0.000 |  |  | 0.940 |
|  | *no* | 86/89 (96.6) | 88/203 (43.4) | reference |  | 10/58 (17.2) | reference |  |
|  | *yes* | 129/130 (99.2) | 72/99 (72.7) | 2.44 [1.61-3.70] |  | 2/11 (18.2) | 0.94 [0.18-4.84] |  |
| Chest pain |  |  |  |  | 0.444 |  |  | 0.018 |
|  | *no* | 150/152 (98.7) | 131/258 (50.8) | reference |  | 7/58 (12.1) | reference |  |
|  | *yes* | 65/67 (97.0) | 29/44 (65.9) | 1.23 [0.72-2.09] |  | 5/11 (45.5) | 5.56 [1.34-23.07] |  |
| Tiredness, exhaustion |  |  |  |  | 0.000 |  |  | 0.001 |
|  | *no* | 31/33 (93.9) | 62/162 (38.3) | reference |  | 3/49 (6.1) | reference |  |
|  | *yes* | 184/186 (98.9) | 98/140 (70.0) | 2.66 [1.79-3.95] |  | 9/20 (45.0) | 11.03 [2.64-45.97] |  |
| Appetite loss |  |  |  |  | 0.000 |  |  | 0.003 |
|  | *no* | 89/93 (95.7) | 104/237 (43.9) | reference |  | 7/61 (11.5) | reference |  |
|  | *yes* | 126/126 (100.0) | 56/65 (86.2) | 4.52 [2.50-8.17] |  | 5/8 (62.5) | 11.30 [2.28-55.89] |  |
| Nausea, vomiting |  |  |  |  | 0.004 |  |  | 0.181 |
|  | *no* | 178/182 (97.8) | 138/275 (50.2) | reference |  | 11/67 (16.4) | reference |  |
|  | *yes* | 37/37 (100.0) | 22/27 (81.5) | 2.93 [1.40-6.13] |  | 1/2 (50.0) | 6.51 [0.42-101.56] |  |
| Diarrhea |  |  |  |  | 0.001 |  |  | 0.475 |
|  | *no* | 154/157 (98.1) | 123/250 (49.2) | reference |  | 10/62 (16.1) | reference |  |
|  | *yes* | 61/62 (98.4) | 37/52 (71.2) | 2.37 [1.41-3.99] |  | 2/7 (28.6) | 1.90 [0.33-11.00] |  |
| Belly pain |  |  |  |  | 0.002 |  |  | 0.530 |
|  | *no* | 175/178 (98.3) | 134/267 (50.2) | reference |  | 10/62 (16.1) | reference |  |
|  | *yes* | 40/41 (97.6) | 26/35 (74.3) | 2.76 [1.44-5.28] |  | 2/7 (28.6) | 1.75 [0.31-10.01] |  |
| Sudden loss of smell or taste |  |  |  |  | 0.000 |  |  | 0.001 |
|  | *no* | 74/77 (96.1) | 96/233 (41.2) | reference |  | 6/62 (9.7) | reference |  |
|  | *yes* | 141/142 (99.3) | 64/69 (92.8) | 6.24 [3.46-11.24] |  | 6/7 (85.7) | 65.25 [5.47-779.10] |  |
| **Use of health services** |  |  |  |  |  |  |  |  |
| Contact with a medical provider |  |  |  |  | 0.000 |  |  | 0.027 |
|  | *no* | 28/28 (100.0) | 86/209 (41.2) | reference |  | 7/57 (12.3) | reference |  |
|  | *yes* | 185/189 (97.9) | 74/93 (79.6) | 3.61 [2.19-5.95] |  | 5/12 (41.7) | 4.62 [1.20-17.86] |  |
| Nasal or throat swabbing to detect SARS-CoV-2 |  |  |  |  | 0.000 |  |  | 0.002 |
|  | *no* | .. (*) | 110/240 (45.8) | reference |  | 7/63 (11.1) | reference |  |
|  | *yes* | .. (*) | 50/62 (80.7) | 2.74 [1.61-4.65] |  | 5/6 (83.3) | 38.52 [3.95-375.53] |  |
| Test result for SARS-CoV-2 † |  |  |  |  | .. |  |  | .. |
|  | *negative or unknown* | .. (*) | 5/16 (31.3) | .. |  | 4/5 (80.0) | .. |  |
|  | *positive* | .. (*) | 45/46 (97.8) | .. |  | 1/1 (100.0) | .. |  |
| Hospitalisation |  |  |  |  | 0.028 |  |  | .. |
|  | *no* | 181/185 (97.8) | 150/291 (51.6) | reference |  | 11/68 (16.2) | .. |  |
|  | *yes* | 32/32 (100.0) | 10/11 (90.9) | 4.96 [1.19-20.62] |  | 1/1 (100.0) | .. |  |
| Admission to the ICU |  |  |  |  | .. |  |  | .. |
|  | *no* | 203/207 (98.1) | 158/300 (52.7) | .. |  | 12/69 (17.4) | .. |  |
|  | *yes* | 10/10 (100.0) | 2/2 (100.0) | .. |  | no participant | .. |  |
| Intubation |  |  |  |  | .. |  |  | .. |
|  | *no* | 210/214 (98.1) | 160/302 (53.0) | .. |  | 12/69 (17.4) | .. |  |
|  | *yes* | 3/3 (100.0) | no participant | .. |  | no participant | .. |  |

All participants, including children and teens. Calculation of odds ratio and p-value: correlation between close contacts of a same index case taken into account using GEE (exchangeable correlation structure, logit link function). \* Index cases all tested positive. † Nucleic acid amplification test.

**Figure 3:**
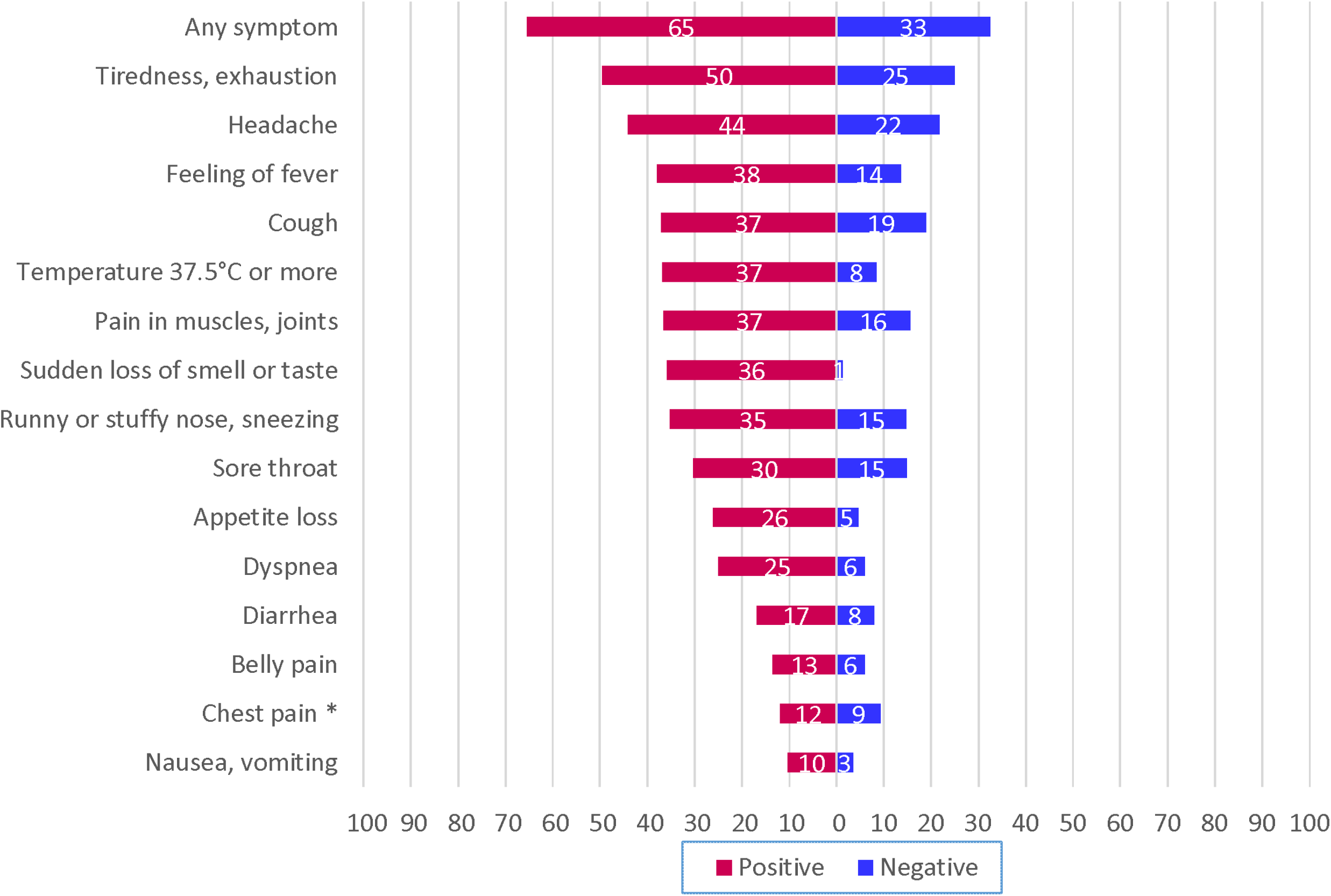
Percentage of household members reporting specific symptoms, according to serology test result. Household members not reporting prior nasal or throat swabbing to detect SARS-CoV-2. Correlation between household members of a same index case taken into account using GEE (exchangeable correlation structure). * Difference not statistically significant at the 0.05 level.

## Discussion

More than one in two participants living with a confirmed COVID-19 case has developed a serologic response to SARS-CoV-2, while one in five close contacts outside the household was seropositive. Our findings confirm that households represent high-risk transmission settings compared with other settings (4,5,8,12,21–23). The SAR we observed is substantially higher than the one reported in previous seroepidemiological studies, including a large nationwide survey conducted in Spain (37·4%), and a retrospective cohort study conducted in Singapore (11%, estimation based on Bayesian modelling) (11–13,23). One study disclosed a SAR of 80% in household members of essential workers, but estimation was based on 30 participants only (10). Beside serological testing characteristics, differences could be due to variable average household sizes (2·2 members in Switzerland vs 2·6 in Spain) (24), unequal adoption of protective behaviours within households (25), or different levels of confinement. Regarding close contacts outside the household, previous seroepidemiological studies provided SAR estimations ranging from 0 to 13·7% (12,14–16,23). Heterogeneity of results could reflect different study designs and settings, and varying adherence to public health protective recommendations (25). The strong difference observed between the prevalence in household members and in close contacts outside home is probably due the fact that contacts at home are closer and last longer than outside, due to the difficulty of applying social distancing in limited spaces and with family members. Simple hygiene rules are theoretically easier to apply at home, but in practice, they tend to be more neglected, maybe due to a feeling of security.

We found that older household members were at particularly high risk, corroborating the findings of previous research on transmission using NAAT (8,26,27). This association was not found for close contacts outside home. This suggests that elderly couples are even less able to apply protective measures at home, due to their high level of mutual dependency. There was no difference in infection susceptibility according to gender, which is in line with other works (5,23,26). The impact of smoking on the risk of SARS-CoV-2 infection is a controversial issue (28). Although household members currently smoking were less frequently tested positive, this association vanished in multivariable analysis, suggesting that it may be confounded by other factors. Our findings suggest that overweight/obesity could lead to increased susceptibility to SARS-CoV-2 infection. The importance of hygiene measures to avoid transmission within household is confirmed by our observations (29). Mask wearing in public and respect of social distancing rules, which is particularly difficult when living under the same roof, were not associated with infection risk in households. In contrast, the association of a greater number of social contacts with a lower probability of infection seems surprising at first place. In fact, our study took place during a period of semi-confinement, during which most people stayed at home, except those who had to go out to work in essential sectors. Our findings thus show that the individual risk of being infected is higher when staying at home than working outside, the aim of confinement (or quarantine) being to break the transmission chain. We have thus to accept that this works well but at the price of a higher risk for household members of COVID-19 cases to be infected. Analogously, the higher probability of infection observed in close contacts outside the household who report always wearing a mask in public (the same trend, although less pronounced, was observed in household contacts) could be due to an association between wearing masks and risk factors for SARS-CoV-2 infection, especially older age. Indeed, during the first epidemic wave in Switzerland, wearing a mask was not yet mandatory and only people at risk tended to use them. Like previous studies, we found an inverse relationship between household size and the proportion of seropositive household members (26,27). This seems counter-intuitive, as prevalence of infectious diseases is well known to be associated with crowded housing. However, in a rich country such as Switzerland, large families can often afford to live in larger apartments, allowing isolation of the sick household member. Being many in a household also allows decreasing mutual dependency and thus close contacts. This association was weakened by inclusion of the mean age of household members in the multivariable model, suggesting that the apparent protective effect of a high number of household members could reflect the fact that large families are, on average younger. However, disentangling respective contributions of household size and age distribution of household members remains difficult.

Regarding the clinical presentation of COVID-19, the proportion of asymptomatic seropositive individuals was close to findings of Pollán and colleagues in Spain (28·5%) (12). Even if not specific, a large number of symptoms were still associated with SARS-CoV-2 infection, especially new-onset smell and/or taste disturbance, confirming the clinical utility of this symptom to suspect COVID-19 (30). Interestingly, the prevalence of flu-like symptoms was high also in seronegative people, maybe because the first epidemic wave occurred just after the winter, when other respiratory infections were still quite prevalent. Moreover, participants knowing that there was a COVID-19 case in their household certainly tended to observe the occurrence of symptoms very closely.

Limitations need to be acknowledged. The Swiss testing policy during the first epidemic wave, which limited diagnostic testing mainly to individuals at increased risk for severe illness, made the sample of index cases not representative of all cases that occurred in the community during this period. Index cases were thus not necessarily the first infected in their household, but those fulfilling testing criteria. However, this would be especially problematic if the purpose were to identify factors associated with infectivity of the index case, which we deliberately avoided.

Incidence of new COVID-19 cases remains high worldwide and prevention of transmission is, for now, the only way to tackle the pandemic. If concerns regarding the transmission of SARS-CoV-2 in shops, restaurants and public gatherings are justified, our findings emphasise that the risk of being infected is much higher at home. In this regard, trying to prevent transmission within households through the dissemination of clear messages on how to protect each other at home is key. However, this remains overlooked in collective awareness and public health discourse, precisely because quarantine and confinement are methods used to break the transmission chain. Early testing of the first case in a household is also important to support immediate self-isolation within the house. Our results suggest in particular that it is essential for non-institutionalised elderly couples to receive strong external support for daily basic needs during the infectious period of the index case. Further research is needed to determine the efficacy and acceptability of specific measures aimed at limiting SARS-CoV-2 transmission within households and at motivating early testing and self-isolation.

## Data Availability

Our data are accessible to researchers upon reasonable request for data sharing to the corresponding author.

## Contributors

JD did the statistical analyses and drafted the first version of the manuscript. JD, MB, SGN and VDA designed the article, accessed the data and contributed to the interpretation of data. MB, SGN and VDA conceived and conducted the study, and contributed to drafting sections of the manuscript. JD, AB, OD, SE, VF, JT, CZ, ME, ASD, SV, MB, SGN and VDA participated in planning of the study and collection of data. GG and GP were involved in development and validation of the serological test. JP and VR provided support for planning and performing statistical analyses. EM contributed to study design. All authors commented on drafts, read and approved the final manuscript. The corresponding author attests that all listed authors meet authorship criteria and that no others meeting the criteria have been omitted.

## Acknowledgments

The authors would like to thank warmly all study participants for their involvement. This study was made possible by the strong involvement of all the *SérocoViD* operational team (Julia Baldwin, Giovanna Bonsembiante-Poidomani, Ophélie Hoffmann, Emilie Jendly, Athiththan Kanthasami, Daria Mapelli, Virginie Schlüter, Kevin Schutzbach, Auriane Soris and Lucie Wuillemin).

The study was funded by the operating budget of the Center for Primary Care and Public Health (Unisanté), University of Lausanne, Switzerland, and by contributions of local health authorities (Department of Health and Social Action, Canton of Vaud) and the following Swiss non-profit institutions: Leenaards Foundation, Fondation pour l’Université de Lausanne. SerocoViD is part of the national Corona Immunitas program coordinated by the Swiss School of Public Health Plus (SSPH+).

---

**Supplementary table 1:** Age distribution and serology test result according to number of participants in the household.

|  | Number of participants in the household, apart from index case |  |  |  |  |
| --- | --- | --- | --- | --- | --- |
|  | 0 | 1 | 2 | 3 | 4 to 6 |
| Number of households (%) | 70/219 (32.0) | 73/219 (33.3) | 30/219 (13.7) | 24/219 (11.0) | 22/219 (10.1) |
| Mean age of all participants, years (SD) | 50.7 (18.8) | 55.8 (16.9) | 40.3 (19.3) | 32.7 (19.7) | 28.3 (17.5) |
| Age of the youngest participant, years | 16.6 | 6.8 | 1.2 | 1.2 | 1.0 |
| Age of the oldest participant, years | 89.1 | 90.1 | 84.8 | 87.3 | 72.6 |
| Mean age of index cases, years (SD) | 50.7 (18.8) | 56.5 (16.5) | 41.6 (21.4) | 41.4 (15.0) | 33.8 (17.8) |
| Age of the youngest index case, years | 16.6 | 17.3 | 2.3 | 9.3 | 4.3 |
| Age of the oldest index case, years | 89.1 | 90.1 | 84.8 | 66.8 | 58.4 |
| Participants with a positive SARS-CoV-2 serology test result, apart from index case, n (%) | .. | 49/73 (67.1) | 32/60 (53.3) | 36/72 (50.0) | 43/97 (44.3) |

**Supplementary table 2:** Serology test result according to symptoms among close contacts not reporting previous nasal or throat swabbing to detect SARS-CoV-2, stratified by type of participant (sensitivity analysis)

|  |  | Household members (N=240) |  |  | Close contacts outside the household (N=63) |  |  |
| --- | --- | --- | --- | --- | --- | --- | --- |
|  |  | n (%) seropositive | Odds ratio [95% CI] | p-value | n (%) seropositive | Odds ratio [95% CI] | p-value |
| Cough | *no* | 68/172 (39.5) | reference | 0.003 | 5/52 (9.6) | reference | 0.308 |
|  | *yes* | 42/68 (61.8) | 2.17 [1.31-3.61] |  | 2/11 (18.2) | 2.46 [0.44-13.80] |  |
| Runny or stuffy nose, sneezing | *no* | 70/180 (38.9) | reference | 0.004 | 5/51 (9.8) | reference | 0.568 |
|  | *yes* | 40/60 (66.7) | 2.24 [1.29-3.86] |  | 2/12 (16.7) | 1.68 [0.28-10.03] |  |
| Sore throat | *no* | 76/186 (40.9) | reference | 0.015 | 7/51 (13.7) | .. | .. |
|  | *yes* | 34/54 (63.0) | 1.89 [1.13-3.15] |  | 0/12 (0.0) | .. |  |
| Dyspnea | *no* | 83/203 (40.9) | reference | 0.000 | 6/57 (10.5) | reference | 0.363 |
|  | *yes* | 27/37 (73.0) | 3.40 [1.74-6.62] |  | 1/6 (16.7) | 2.59 [0.33-20.04] |  |
| Feeling of fever | *no* | 67/179 (37.4) | reference | 0.000 | 6/51 (11.8) | reference | 0.915 |
|  | *yes* | 43/61 (70.5) | 2.99 [1.67-5.34] |  | 1/12 (8.3) | 0.89 [0.11-7.04] |  |
| Temperature 37.5°C or more (measured) | *no* | 69/187 (36.9) | reference | 0.000 | 7/61 (11.5) | .. | .. |
|  | *yes* | 41/53 (77.4) | 4.04 [2.21-7.39] |  | 0/2 (0.0) | .. |  |
| Headache | *no* | 60/159 (37.7) | reference | 0.004 | 4/51 (7.8) | reference | 0.080 |
|  | *yes* | 50/81 (61.7) | 2.01 [1.24-3.24] |  | 3/12 (25.0) | 4.29 [0.84-21.84] |  |
| Pain in muscles, joints | *no* | 70/178 (39.3) | reference | 0.003 | 7/55 (12.7) | .. | .. |
|  | *yes* | 40/62 (64.5) | 2.14 [1.29-3.56] |  | 0/8 (0.0) | .. |  |
| Chest pain | *no* | 97/214 (45.3) | reference | 0.737 | 5/56 (8.9) | reference | 0.064 |
|  | *yes* | 13/26 (50.0) | 0.89 [0.44-1.78] |  | 2/7 (28.6) | 5.37 [0.91-31.75] |  |
| Tiredness, exhaustion | *no* | 56/153 (36.6) | reference | 0.001 | 3/49 (6.1) | reference | 0.023 |
|  | *yes* | 54/87 (62.1) | 2.32 [1.45-3.73] |  | 4/14 (28.6) | 6.70 [1.30-34.51] |  |
| Appetite loss | *no* | 82/206 (39.8) | reference | 0.000 | 4/57 (7.0) | reference | 0.005 |
|  | *yes* | 28/34 (82.4) | 5.01 [2.30-10.91] |  | 3/6 (50.0) | 14.46 [2.22-94.35] |  |
| Nausea, vomiting | *no* | 98/224 (43.8) | reference | 0.038 | 6/62 (9.7) | .. | .. |
|  | *yes* | 12/16 (75.0) | 2.60 [1.06-6.41] |  | 1/1 (100.0) | .. |  |
| Diarrhea | *no* | 91/209 (43.5) | reference | 0.013 | 6/57 (10.5) | reference | 0.629 |
|  | *yes* | 19/31 (61.3) | 2.33 [1.20-4.53] |  | 1/6 (16.7) | 1.76 [0.18-17.28] |  |
| Stomach ache | *no* | 95/217 (43.8) | reference | 0.009 | 6/57 (10.5) | reference | 0.457 |
|  | *yes* | 15/23 (65.2) | 2.76 [1.29-5.90] |  | 1/6 (16.7) | 2.28 [0.26-19.98] |  |
| Sudden loss of smell or taste | *no* | 71/198 (35.9) | reference | 0.000 | 3/58 (5.2) | reference | 0.001 |
|  | *yes* | 39/42 (92.9) | 9.81 [4.44-21.68] |  | 4/5 (80.0) | 71.91 [6.19-835.19] |  |

All participants, including children and teens. Calculation of odds ratio and p-value: correlation between close contacts of a same index case taken into account using GEE (exchangeable correlation structure, logit link function).

