## Supplementary material for "Prevalence of SARS-CoV-2 in household members and other close contacts of COVID-19 cases: a serologic study in canton of Vaud, Switzerland"

**Supplementary material: e-Questionnaire filled by adult close contacts within household and outside (versions filled by index cases and child/teen participants are available upon request)**

unisanté
Centre universitaire de médecine générale et santé publique · Lausanne

### SérocoViD

#### A. Personal Data

| Variable | Type | Réponses |
|---|---|---|
| 1. Last name | Texte | |
| 2. First name | Texte | |
| 3. Date of birth | Date | dd/mm/yyyy |
| 4. Gender | Catégoriel | Woman / man / other |
| *Home address* | | |
| 5. Care of (C/o) | Texte | |
| 6. Street name | Texte | |
| 7. Street number | Nombre | ### |
| 8. Floor number | Nombre | ## |
| 9. Postcode | Nombre | #### |
| 10. Locality (City/Town/Village) | Texte | |
| 11. Phone number | Nombre | ### / ### ## ## |
| 12. Email address | Texte | __________@_____________ |

#### B. Questionnaire data

| Variable | Type | Réponses |
|---|---|---|
| 13. I am filling in this questionnaire for | Catégoriel | myself / other person |
| 14. Date of filling in the questionnaire | Date | dd/mm/yyyy |
| 15. Where are you filling in the questionnaire (only one possible answer)? | Menu déroulant | Home or work / Lausanne study center / Rennaz study center / Yverdon study center/ Nyon study center |
| 16. I am filling in this questionnaire with the help of a study investigator | Binaire | yes / no |
| 17. *Si oui à 16.* Last name of the study investigator | Texte | |
| 18. *Si oui à 16.* First name of the study investigator | Texte | |

#### C. Medical history

| Variable | Type | Réponses |
|---|---|---|
| 19. *Si femme à 4.* Are you pregnant or have you given birth in 2020? | Categoriel | no / I am currently pregnant/ I gave birth in 2020 |
| 20. *Si enceinte à 19.* When is your due date? | Date | dd/mm/yyyy |
| 21. *Si accouché à 19.* When did you give birth? | Date | dd/mm/yyyy |
| Do you have the following diseases? | | |
| 22. Arterial hypertension (high blood pressure) | Binaire | yes / no |
| 23. *Si oui à 22.* Does this hypertension require medication? | Binaire | yes / no |
| 24. *Si oui à 23.* Is this hypertension well controlled with the medication? | Binaire | yes / no |
| 25. Diabetes | Binaire | yes / no |
| 26. *Si oui à 25.* Does this diabetes require medication? | Binaire | yes / no |
| 27. *Si oui à 25.* Is this diabetes well controlled with this medication? | Binaire | yes / no |
| 28. Cardiovascular disease (heart failure, history of heart attack or stroke, heart valve disease) | Binaire | yes / no |
| 29. Kidney disease (impaired renal function) ? | Binaire | yes / no |
| 30. Chronic respiratory disease (other than asthma well controlled by medication and sleep apnea syndrome) | Binaire | yes / no |
| 31. Immune system weakness due to a disease or a treatment | Binaire | yes / no |
| 32. *Si oui à 28.* Which disease or treatment? | Texte | |
| 33. Cancer currently under treatment | Binaire | yes / no |
| 34. Other chronic disease(s) or recurrent health problem | Binaire | yes / no |
| 35. *Si oui à 31.* Which disease? | Texte | |
| 36. Do you currently take any medication? | Binaire | yes / no |
| 37. *Si oui à 33.* List all the medications you are taking (including painkillers, tranquilisers, sleeping pills and natural remedies) | Texte | Saisie semi-automatique |
| *38.* Currently, do you smoke (even occasionally) or use products containing nicotine (including electronic cigarettes and iQOS)? | Catégoriel | yes / no |
| *39. Si oui à 38.* Currently, how often do smoke or use these products ? | Catégoriel | daily / at least once a week but not everyday / less than once a week |
| *40. Si oui à 38.* What do you smoke / which product do you use (several answers possible) ? | Menu déroulant | - cigarettes<br>- electronic cigarettes (vaping) with or without nicotine<br>- heated tobacco products like iQOS (Pax, etc.)<br>- nicotine substitutes (patches, gums, inhaler,…)<br>- other |
| 41. *Si des cigarettes à 40.* On average, how many cigarettes do you smoke per day (less than one cigarette = 0, 1 pack = 20 cigarettes, / half a pack = 10 cigarettes) | Nombre | ## |

| | | |
|---|---|---|
| 42. *Si des cigarettes électroniques à 40.* Which dosage of nicotine do you use on average (mg/ml) ? | Catégoriel | 0 (sans nicotine) / moins de 6 / 6-12 / 13 ou plus |
| 43. *Si non à 38.* Have you ever smoked regularly for more than 6 months ? | Binaire | yes / no |
| 44. *Si oui à 43.* When did you quit smoking ? | Catégoriel | - In the past 12 months<br>- Between one year and less than 2 years ago<br>- Between 2 years and less than 5 years ago<br>- Between 5 years and less than 10 years ago<br>- 10 years ago or more |
| 45. *Si oui à 38 ou oui à 43.* At what age did you start smoking regularly? | Nombre | ## |
| 46. How tall are you (cm)? | Nombre | ### |
| 47. How much do you weigh (kg)? | Nombre | ###.# |
| 48. Have you gained weight since the end of February 2020? | Binaire | yes / no |
| 49. *Si oui à 43.* How many kilos? | Nombre | ## |
| 50. Have you lost weight since the end of February 2020? | Binaire | yes / no |
| 51. *Si oui à 45.* How many kilos? | Nombre | ## |
| 52. Do you get vaccinated against the flu? | Catégoriel | Yes, every year (or almost), this winter included / yes, only this winter / yes but not this winter / no / I don't know |
| 53. Have you ever been vaccinated against tuberculosis (BCG vaccine, the one that sometimes leaves a mark)? | Binaire | yes / no / I don't know |
| 54. Since the end of February 2020, how many flu episodes (respiratory symptoms or impression of fever or sudden loss of taste/smell) did you have (without link to chronic disease or already known allergy) ? | Catégoriel | 0 / 1 / 2 / 3 / 4 / 5 |

#### D. Flu-like symptoms and utilisation of health services *(sous-questionnaire si oui à 47, un par épisode)*

| Variable | Type | Réponses |
|---|---|---|
| 1. When did this epsiode of flu-like symptoms begin? *(even if you are not totally sure of the date, please provide a most probable one)* | Date | dd/mm/yyyy |
| 2. Is this episode finished? | Binaire | yes / no |
| 3. *Si oui à 2.* When did this episode finish (end of main symptoms)? | Date | dd/mm/yyyy |
| During this this episode of flu-like symptoms? (several possible answers) | | |
| 4. I coughed | Coche | |
| 5. I had a runny or stuffy nose, I sneezed | Coche | |
| 6. I had a sore throat | Coche | |
| 7. I was out of breath | Coche | |
| 8. It felt like I had a fever | Coche | |
| 9. I had a temperature of 37.5°C or higher (measured) | Coche | |
| 10. I had a headache | Coche | |
| 11. I had aching muscles / joints | Coche | |
| 12. I had chest / thorax/ sternum pain | Coche | |
| 13. I felt tired, exhausted | Coche | |
| 14. I lost my appetite | Coche | |
| 15. I felt nauseous, I was sick (vomited) | Coche | |
| 16. I had diarrhea | Coche | |
| 17. I had a stomach ache | Coche | |
| 18. All of a sudden, I lost my sense of smell or of taste for food/beverages | Coche | |
| 19. Other | Coche | |
| 20. *Si coche à 18.* What other symptoms did you have? | Texte | |
| 21. Have you taken any anti-inflammatory drug such as aspirin (Aspégic), ibuprofen (Algifor, Irfen, Brufen) or diclofenac (Voltaren) or any true anti-inflammatory drug to treat your symptoms? | Binaire | yes / no |
| 22. Did you contact or go to a healthcare provider (medical practice, emergency, telephone center) for this episode? | Binaire | yes / no |
| *Si oui à 21*. Which healthcare provider(s)? (several possible answers) | | |
| 23. hotline | Coche | |
| 24. On-call doctors telephone center (Centrale Téléphonique des Médecins de garde - CTMG) | Coche | |
| 25. Ambulance (144) | Coche | |
| 26. General Practitioner (GP) | Coche | |
| 27. Pharmacist | Coche | |
| 28. Medical centre or outpatient clinic | Coche | |
| 29. Si oui à 26. Which one? | Texte | |
| 30. Other healthcare provider? | Texte | |

| | | |
|---|---|---|
| 31. Were you admitted to hospital? | Binaire | yes / no |
| 32. *Si oui à 29.* In which establishment? | Texte | |
| 33. *Si oui à 29.* When were you admited to the hospital? | Date | dd/mm/yyyy |
| 34. *Si oui à 29.* When were you discharged from hospital? | Date | dd/mm/yyyy |
| 35. *Si oui à 29.* Were you admitted to the Intensive Care Unit (ICU) during this hospitalisation? | Binaire | yes / no |
| 36. *Si oui à 33.* Were you intubated during this hospitalisation? | Binaire | yes / no |
| 37. Were you tested for COVID-19 by nasal or throat swab during this episode of flu-like symptoms? | Binaire | yes / no |
| 38. *Si oui à 35.* What were the results of this test(s) (several possible answers)? | Menu déroulant | positive / negative / first negative than positive / still waiting for the results / don't know |
| 39. Did you got to work during this episode of flu-like symptoms? | Binaire | yes / no |
| 40. *Si oui à 37.* For how many days in total? | Nombre | ### |
| 41. Did you stay at home without going out at all during this episode? | Binaire | yes / no |
| 42. *Si oui à 39.* For how many days in total? | Nombre | ### |
| 43. Among the people living under the same roof as you, how many of them stayed at home without going out at all? | Nombre | # |
| 44. *Si ≥1 à 41.* For how many days in total?<br>Person 1<br>Person 2<br>Person 3… | <br>Nombre<br>Nombre<br>Nombre… | <br>##<br>##<br>##... |
| Which preventive measures did you take at home during this episode? (several possible answers) | | |
| 45. You had food or other essentiel products, such as medecine, delivered to your house by a family member, a friend or a delivery service | Coche | |
| 46. You stayed in one room alone, with the door closed, and ate inside the room | Coche | |
| 47. You aired the room several times a day | Coche | |
| 48. You avoided all visitors and contact | Coche | |
| 49. You only left the room if necessary | Coche | |
| 50. You had a trash can in the room dedicated to used tissues | Coche | |
| 51. You avoided all contact with pets | Coche | |
| 52. You used a bathroom solely dedicated to you, or if not possible, you cleaned the bathroom (sink, toilets, shower, bathtub) after each use with a regular household disinfectant | Coche | |
| 53. You washed your hands before preparing food | Coche | |
| 54. You did not share cutlery, glasses, cups and kitchen utensils and you washed them carefully | Coche | |
| 55. You washed clothes, bedding and towels several times a week | Coche | |
| 56. You wore a mask when you left the room | Coche | |
| 57. You asked every person that entered the room to wear a mask | Coche | |
| 58. You cleaned and disinfected, daily, any surface that you touched (door handles, bed frames and other bedroom furniture) with a regular household disinfectant | Coche | |

#### E. Place where you live

| Variable | Type | Réponses |
|---|---|---|
| 55. What type of home do you live in? | Catégoriel | Individual house / apartment / studio / residential home / other |
| 56. *Si autre à 55,* Please specify | Texte | |
| 57. What is the approximate surface area of your home (m2)? | Nombre | ### |
| 58. How many habitable rooms do you have (apart from the kitchen)? | Nombre | ## |
| 59. How many separate bathrooms or toilets does your home have? | Nombre | # |
| 60. Do you have a balcony or a terrace? | Binaire | yes / no |
| 61. Do you have a private or shared garden? | Binaire | yes / no |
| 62. Do you have pets? | Catégoriel | dog / cat / dog and cat / other / none |
| 63. Since the end of February 2020, how many people live under the same roof as you (i.e. people sleeping there at least 3 nights per month)? | Nombre | ## |

#### F. Behaviour and exposure (in private)

| Variable | Type | Réponses |
|---|---|---|
| Today, in your <u>private</u> life, would you say that you are following proper preventive measures and practices to protect yourself and others from coronavirus, such as: | | |
| 64. Following simple hygiene rules (wash hands regularly, sneeze into your elbow, use a disposable tissue, etc.)? | Catégoriel | yes / mostly yes / mostly no / no |

| | | |
|---|---|---|
| 65. Respecting « social distancing » rules (avoid shaking hands or kissing, stay at home, avoid leaving your home unless absolutely necessary, etc.)? | Catégoriel | yes / mostly yes / mostly no / no |
| 66. During the confinement (March 16 to May 10), on average, how many times per week do you or did you go outside to do grocery shopping? | Nombre | ## |
| 67. *Si >0 à 66.* Where do you do your grocery shopping? | Binaire | Mainly small nearby businesses/ mainly supermarkets / both |
| 68. *Si >0 à 68.* What mode of transport do/did you use most of the time to do your grocery shopping (only one possible answer)? | Menu déroulant | car / public transport / motorcycle / bike / scooter / on foot |
| 69. *Si >0 à 68.* What other mode of transport (second most frequent) do/did you use to do your grocery shopping (only one possible answer)? | Menu déroulant | car / public transport / motorcycle / bike / scooter / on foot / no other |
| 70. During the confinement (March 16 to May 10), on average, how many times per week did other people living under the same roof as you go/went out to do grocery shopping ? | Nombre | ## |
| During the confinement (March 16 to May 10), which other trips did you make? | | |
| 71. A trip to go to your work place (number of times) | Nombre | ### |
| 72. A trip to go to the pharmacy (number of times) | Nombre | ## |
| 73. A trip to go to the doctor (number of times) | Nombre | ## |
| 74. A trip to go to a therapist other than a doctor (number of times) | Nombre | ## |
| 75. A trip to go to the hairdresser or any other beauty care provider (number of times) | Nombre | ## |
| 76. A trip to go to a garden center or hardware store (number of times) | Nombre | ## |
| 77. A trip to help your relatives (number of times) | Nombre | ## |
| 78. A trip to visit people living in a EMS (seniors residential home) (number of times) | Nombre | ## |
| 79. A trip to visit family (number of times) | Nombre | ## |
| 80. A trip to visit friends (number of times) | Nombre | ## |
| 81. Other visits or trips (type) | Texte | |
| 82. Other trips or visits (number of times) | Nombre | ## |
| 83. During the confinement (March 16 to May 10), what mode of transport do/did you use most of the time for these trips (only one possible answer)? | Menu déroulant | car / public transport / motorcycle / bike / scooter / on foot |
| 84. During the confinement (March 16 to May 10), what other mode of transport (second most frequent) do/did you use for these trips (only one possible answer)? | Menu déroulant | car / public transport / motorcycle / bike / scooter / on foot / no other |
| 85. During the confinement (March 16 to May 10), on average, how many people do/did you meet per week apart from the people living under the same roof as you? | Catégoriel | 0 / 1 to 2 / 3 ot 5 / 6 ot 10 / more than 10 |
| *Si >0 à 78.* In what context are/were you meeting these people? | | |
| 86. Meetings with friends | Coche | |
| 87. Family reunions | Coche | |
| 88. Work | Coche | |
| 89. Public transport | Coche | |
| 90. By car | Coche | |
| 91. Sports | Coche | |
| 92. Other | Coche | |
| 93. *Si coche à 85.* Please specify | Texte | |
| 94. During the confinement (March 16 to May 10), did you reduce the number of people you normally met/meet? | Catégoriel | Yes a lot / yes moderately / no |
| 95. Do you wear a mask in public? | Catégoriel | no / yes, sometimes / yes, all the time |
| *Si oui, parfois à 88.* In which situations (several possible answers) ? | | |
| 96. In public transport | Coche | |
| 97. In shops | Coche | |
| 98. Other | Coche | |
| 99. *Si coche à 91.* Please specify | Texte | |
| *Si oui, parfois / oui, toujours à 88.* Which type of mask do/did you use in public spaces (several possible answers)? | | |
| 100. Simple medical mask (also known as a surgical mask) | Coche | |
| 101. Disposable mask made by myself | Coche | |
| 102. Respiratory protection mask (also know as shell, duck or FFP2 mask) | Coche | |
| 103. Cloth mask | Coche | |
| 104. Did the confinement influence your consumption of cigarettes or other products with nicotine? | Menu déroulant | - I stopped smoking / using these products<br>- I diminished my consumption<br>- I did not change my consumption<br>- I increased my consumption<br>- I started again<br>- I did not start again<br>- I have never smoked / used these products |

| | | |
|---|---|---|
| 105. Since the end of February 2020, how many times have you travelled outside of Switzerland (apart from a professional activity in a neighbouring area)? | Nombre | ## |
| *Si ≥1 à 97. Pour chaque voyage:* | | |
| 106. In which country (several possible answers)? | Menu déroulant | All the countries |
| 107. Start date | Date | dd/mm/yyyy |
| 108. End date | Date | dd/mm/yyyy |
| 109. Apart from the people living under the same roof as you, how many people were you in close contact with (at less than 2 meters for more than 15 minutes) who had symptoms suggestive of COVID-19 (fever or cough or fatigue or out of breath or muscular pain or loss of taste/smell) while they were sick (or 48 hours before they were sick)? | Nombre | ## |
| *Si ≥1 à 101. Pour chaque personne :* | | |
| 110. What is the gender of this person? | Catégoriel | woman / man / other |
| 111. How old is this person? | Nombre | ## |
| 112. When was this contact made? *(even if you are not totally sure of the date, please provide a most probable one)* | Date | dd/mm/yyyy |
| 113. Was this person tested for COVID-19 with a nasal or throat swab during this episode? | Binaire | yes / no / don't know |
| 114. *Si oui à 105.* What were the results of the test(s) (only one possible answer)? | Menu déroulant | Positive / negative/ first negative than positive / still waiting for the results / I don't know |

#### G. Formation, situation professionnelle

| **Variable** | **Type** | **Réponses** |
|---|---|---|
| 115. What is the highest level of education that you have succesfully achieved and for which you have obtained a diploma or a certificate (only one possible answer)? | Menu déroulant | - No diploma<br>- Compulsory school (Certificate of end of secondary school)<br>- General/ vocational secondary education (Trade school, diploma)<br>- Secondary education preparing for a trade (CFC)<br>- General secondary education, superior level (maturité)<br>- General secondary education preparing for a trade, superior level (specialised maturité)<br>- Non-university higher education, 3 years or less<br>- Non-university higher education, 3 years or more<br>- University, EPFL/EPFZ |
| 116. Which category best corresponds to your professional activity during the greatest number of years (only one possible answer)? | Menu déroulant | - Labourer, worker<br>- Skilled worker, foreman<br>- Farmer<br>- Unqualified employee (e.g. Office assistant)<br>- Qualified employee (e.g. secretary, accountamt)<br>- Middle management (e.g. technician, teacher)<br>- Independant small buisness owner, craftsman<br>- Senior management (e.g. economist, company lawyer)<br>- Liberal profession (e.g. doctor, lawyer)<br>- Director, head of a company or of a public service<br>- At home without a profession, retired or no longer in business<br>- Did not work for health reasons (invalidity, chronic disease, …)<br>- Did not work for reasons other than health (e.g. student) |
| Currently, you are? | | |
| 117. Retired | Coche | |
| 118. A student | Coche | |
| 119. An independent worker | Coche | |
| 120. An employee | Coche | |
| 121. A homemaker (house-wife or house-husband) | Coche | |
| 122. Unemployed | Coche | |
| 123. Other | Coche | |

| | |
|---|---|
| 124. *Si coche à 115.* Please specify | Texte |

#### H. Exposure at work *(sous-questionnaire si coche à 118 ou 119 ou 122)*

| Variable | Type | Réponses |
|---|---|---|
| 125. What is your profession? | Texte | |
| In which sector(s) do you study or work? (several possible answers) | | |
| 126. Agriculture | Coche | |
| 127. Industrial manufacturing | Coche | |
| 128. Production and distribution of energy, of water | Coche | |
| 129. Waste management | Coche | |
| 130. Construction | Coche | |
| 131. Agri-food trade | Coche | |
| 132. Whole sale and retail sale | Coche | |
| 133. Transportation and storage | Coche | |
| 134. Postal and courier activities | Coche | |
| 135. Accomodation and catering | Coche | |
| 136. Information and communication | Coche | |
| 137. Financial and insurance activities, real estate | Coche | |
| 138. Specialised, scientific and technical activities | Coche | |
| 139. Legal, accounting and management activities | Coche | |
| 140. Architectural and engineering acitivities | Coche | |
| 141. Scientific research and development | Coche | |
| 142. Administrative and support service activities | Coche | |
| 143. Public administration | Coche | |
| 144. Education | Coche | |
| 145. Human health and social action | Coche | |
| 146. Arts, entertainment and recreation | Coche | |
| 147. Other | Coche | |
| 148. *Si coche à 140.* Please specify | Texte | |
| 149. *Si coche à 138.* In what type of establishment do you work (only one possible answer)? | Menu déroulant | hospital / outpatient clinic / medical office / EMS / ESE / CMS |
| 150. *Si coche à 148.* During your work, are you or were you in direct contact with sick people? | Binaire | yes / no |
| *151. Si oui à 148.* Among these sick patients, are there any that are or were suspected of having COVID-19? | Binaire | yes / no / don't know |
| 152. *Si coche à 138.* During your work, did you wear a mask (surgical mask, II or or IIR)? | Catégoriel | Yes, all the time / yes, only when I was in contact with a sick person / no |
| 153. *Si coche à 138.* Was there a period of time during which you were in contact with a sick person and that you were unable to wear a mask? | Binaire | yes / no |
| 154. *Si oui à 145.* During how many days? | Nombre | ## |
| 155. Since the beginning of the COVID-19 epidemic, has there been a change in your working conditions? | Binaire | yes / no |
| *Si oui à 147.* Please specify (several possible answers) | | |
| 156. *Si oui à 147.* Decrease in activity | Coche | |
| 157. *Si oui à 147.* Stopped activity | Coche | |
| 158. *Si oui à 147.* Use of teleworking (remote working) | Coche | |
| 159. *Si oui à 147.* Sick leave | Coche | |
| 160. *Si oui à 147.* Unemployment | Coche | |
| 161. *Si oui à 147.* Other | Coche | |
| 162. *Si coche à 153.* Please specify | Texte | |
| 163. *Si pas de coche à 148.* During the confinement (March 16 to May 10) did you continue to go to your workplace? | Menu déroulant | yes most of the time / yes, but only part of the time / no |
| 164. *Si oui à 166,* In relation to the preventive measures recommended by the authorities? | | |
| 165. Were you able to respect the hygiene rules at work? | Menu déroulant | yes / mostly yes / mostly no / no |
| 166. Were you able to respect the « social distancing » measures at work? | Menu déroulant | yes / mostly yes / mostly no / no |
| 167. Was there disinfectant made available at work? | Binaire | yes / no |
| 168. Were masks available at work? | Binaire | yes / no |
| 169. Were physical barriers (i.e. plexiglas) made available at work? | Binaire | yes / no |
| 170. Since the beginning of the COVID-19 epidemic, did you go to work at the Swiss border or on the other side of the border? | Binaire | yes / only in the beginning / no |
| 171. *Si oui à 167.* Please specify in which neighbouring country/countries you went to work (several possible answers) | Menu déroulant | France / Italy / Germany / Austria / Liechtenstein |
